# Lower delayed but comparable working memory performance in patients with Parkinson disease

**DOI:** 10.1101/2023.03.31.23288016

**Authors:** Eun-Young Lee

## Abstract

**Intruduction:** The present study examined nature of memory deficits and its associations with MRI structural metrics in patients with Parkinson disease (PD).

**Methods:** Nineteen PD and 23 matched controls underwent two memory experiments. In experiment 1 (delayed memory task), subjects were asked to remember an array of colored rectangles with varying memory set sizes [Low-Load (2 items), Low-Load with distractors, & High-Load (5 items)]. After a 7s delay period, they reported whether the orientation of any relevant figures had changed (test period). In experiment 2 (working memory task), memory arrays were presented in varying set sizes (2 to 6 items) but with no distractors and they were followed by a 2s delay period and subsequent test period. Brain MRI data were acquired to assess morphological differences (volumes and cortical thickness) in brain areas related to attentional filtering (middle frontal gyrus and basal ganglia) and memory storage and consolidation (intraparietal sulcus and medial temporal corticies).

**Results:** Compared to controls, PD patients had lower memory capacity scores in all memory conditions for the experiment 1 (p <0.021) whereas there were no memory score group differences in any memory set sizes for the experiment 2 (p>0.06). In addition, there were no group differences in structural metrics for any ROI and no asccociations of strucrutal metrics with delayed memory scores (p>0.056).

**Discussion:** The present findings suggest that both lower attentional filtering and memory storage may contribute to lower delayed memory scores in PD. Memory storage capacity for visuospatial working memory in PD, however, appeared to be comparable to that of controls at least in the absence of apparent distractors. In addition, lower delayed memory in PD may partly be associated with early brain structural or functional changes that may occur before morphological changes.

## 1. Introduction

Parkinson’s disease (PD) is a slowly progressing neurodegenerative disorder with predominant loss of dopaminergic neurons in SNc (subtantia nigra pars compacta) and subsequent depletion of dopamine levels in the basal ganglia. Prominent characteristics of Parkinson’s disease include motor symptoms such as tremor, rigidity and bradykinesia. While the motor symptoms of PD dominate the clinical picture, many patients with PD experience a wide range of non-motor symptoms. These may include autonomic disturbances (e.g., constipation and bladder control problems) and sensory complaints (e.g., numbness, burning or tingling sensation) but also include psychiatric (e.g., depression and anxiety) or non-psychiatric cognitive dysfunctions that may represent difficultyies with memory and attention (Gabrieli et al., 1996; Zgaljardic et al., 2003; Owen, 2004).

Given that the basal ganglia have extensive interconnections with the prefrontal cortex (Jellinger, 2001; Lewis et al., 2003), patients’ cognitive symptoms are often ascribed to compromised information flow through this pathway (Lewis et al., 2003). In fact, the pattern of cognitive deficits in patients with Parkinson’s disease appears to be similar to that observed in frontal lobe patients such as planning, selective attention, and set shifting (Morris et al., 1988, Owen, 1995; West et al., 1998). Converging evidence, however, suggests that PD may experience various cognitive dysfunctions beyond frontal dysfunctions (e.g., learning new information and holding the information for longer period of time in memory, visuospatial perception, and language disturbunces that are typically considered as functions related to posterior regions of the brain) since more than 80% PD patient may convert to dementia within 20 years of their PD diagnosis.

In the present study, we examined whether lower memory performance in PD may partly be due to reduced storage capacity per se (related to posterior lobe dysfunction) or inability to filter out irrelevant information (related to frontal lobe dysfunction), or both. Our central hypotheses were: 1) compared to controls, PD patients will show lower performance in both working memory and delayed memory particularly for the high load conditions 2) The memory performance in PD will become lower when distractors were presented compared to without distractors; 3) There will be significant morphological differences that were assessed by volume or cortical thickness particularly in the areas related to attentional filtering (e.g., middle frontal gyrus and basal ganglia) (McNab and Klingberg, 2008) and memory (working memory or delayed memory) storage (intraparietal sulcus and medial temporal lobe areas); 4) The MRI structural metrics (volume and cortical thickness) in ROIs will be associated with memory performance metrics.

## 2. Materials and Methods

### 2.1. Participants

Fourty two subjects (19 patients with Parkinson’s disease (PD) and 23 age- and education-matched neurologically normal subjects) were recruited for the study. All subjects except for one patient and one control subject reported having normal color vision and normal or corrected-to-normal acuity. One patient and one control who were identical twins were partially color-blind, but they were able to tell the difference between red and green rectangles used in this study. All subjects had Mini-Mental Status Examination (MMSE) scores >=26. Depression was evaluated by means of Geriatric Depression Scale-long form (GDS; Sheikh and Yesavage, 1986). Three patients and one control subject were taking antidepressants (e.g., Fluoxetine, Bupropion, or Sertraline) at the time of the study. The general pattern of results was essentially the same with or without these three patients, so their data have been retained.

PD patients were free from other neurological disorders. Fifteen patients were receiving the dopamine precursor Levodopa as treatment. On patient was not taking any anti-parkinsonian medication. Another patient was taking only Pramipexole (a dopamine agonist). The remaining two patients were receiving Ropinirole in conjunction with Trihexyphenidyl (an anticholinergic agent) or Azilect (an MAO inhibitor). On the morning of the experiment, PD patients skipped their initial dose of anti-parkinsonian medication. The mean withdrawal period of 11 h (at least 9 h) would not be enough to achieve complete clearance. Rather, it was intended to enhance differences between experimental groups while minimizing the burden imposed on patients. The severity of the disease was reassessed just before the start of the experiment using the Hoehn and Yahr scale (1967). Control subjects reported neither a history of neurological problems nor any significant current psychiatric disorder. All participants gave their informed consent according to procedures approved by the ethics board at the University of Missouri-Columbia (Approval number: 1170557).

### 2.2. Stimuli and Procedures for Neuropsychological Experiments

Stimulus arrays were presented within a 4 × 7.3° rectangular region centered at fixation on a dark background. Arrays consisted of either two or five colored rectangles. Item positions were randomized across trials. Both red and green rectangles subtended .65 × 1.15° of visual angle, with orientations selected randomly from a set of four possible values (vertical, horizontal, left-tilting 45°, and right-tilting 45°).

In experiment 1 (Figure 1), each trial began with a 2 s get-ready signal (3 yellow crosses). Next there was an instructional cue, which was followed by a 1 s long memory array, consisting of either two red or two red and three green rectangles. The instructional cue indicated whether subjects should ignore the green rectangles as distracters (“**X**”: Low Load+ Distracter) or remember them as part of target memory array with no distracters (“**o**”: Low Load, or “**O**”: High Load).

**Figure 1.**
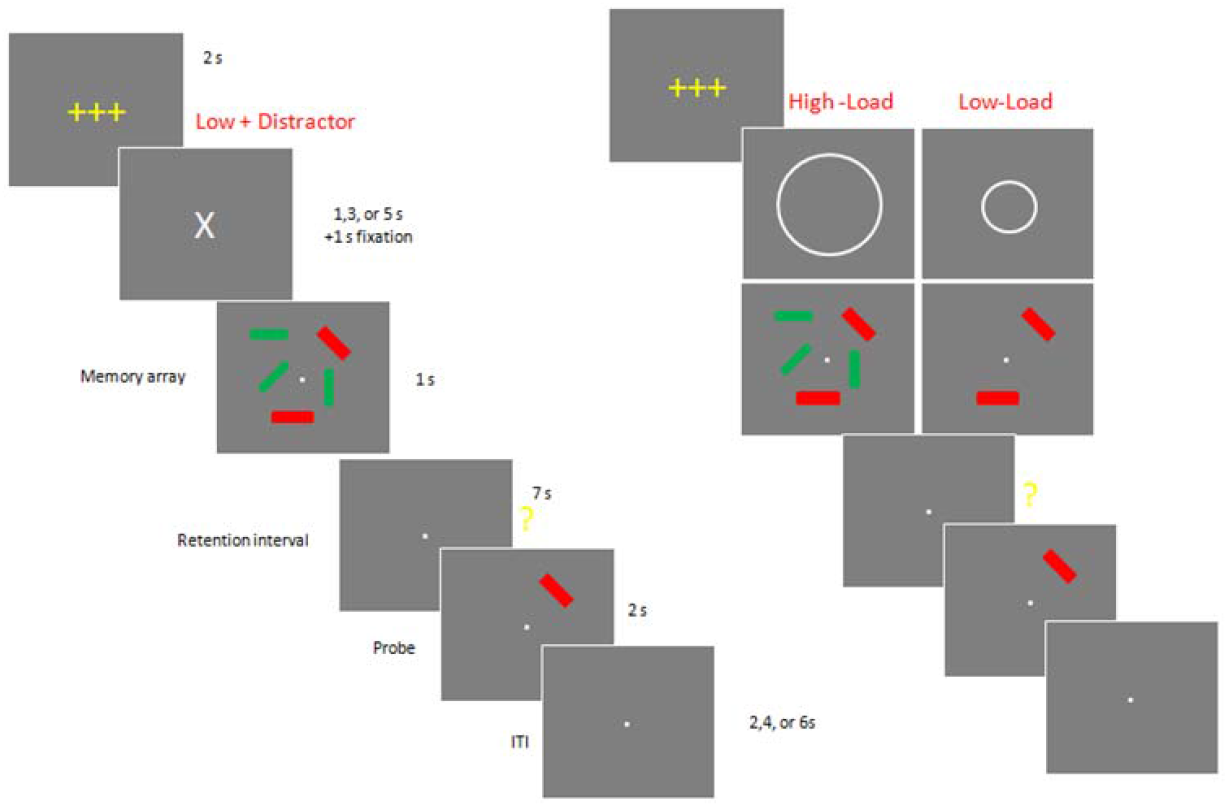
Example of a typical trials of Experiment 1

After a 7-s long retention interval, subjects were presented with a single probe stimulus for 2 s and asked to press a specified button on either the left- or right-hand key pad to report whether the orientation of the tested rectangle changed (same or different). Following an inter-trial interval of 2, 4 or 6 s, the next trial commenced, starting with the yellow get-ready signal. Accuracy was emphasized over speed, and subjects were allowed to correct their response before the next trial began. The experiment 2 consists of twelve blocks of 12 trials (total 144 trials). Between blocks, participants were allowed to take as long a break as they wanted.

In experiment 2, subjects performed a change detection task to estimate their working memory capacity. Subjects sat upright in a comfortable chair and viewed the stimuli at a distance of ∼70 cm. In this version of the task, there were neither precues nor green distracters, only relevant items. The number of to-be-remembered red rectangles varied from 2 to a maximum of 6, which slightly exceeds the typical capacity of an older adult. Each trial began with a 2 s get-ready signal followed by a 1 s long memory array. After a brief (200 ms) pattern mask and a 2 s retention interval, the test stimulus was presented until a response was made. Following a 3 s inter-trial interval, the next trial commenced. Accuracy was emphasized over speed, and subjects were allowed to correct their response before the next trial began. This version of the task was structured as five blocks of 32 trials (total 160 trials).

### 2.3. MRI image acquisition and image processing

Images were acquired on the 3-Tesla Siemens scanner at the University of Missouri’s Brain Imaging Center. The session began with a T1-weighted structural scan, upon which the functional images were ultimately co-registered. Technical parameters for the structural scans were as follows: T1-weighted MPRAGE images: repetition time (TR) =1920 ms, echo time (TE) =2.92 ms, flip angle=9º, field of view (FOV) = 256 mm, matrix: 256 × 256, 176 slices in the sagittal plane, voxel size = 1×1×1 mm, slice thickness = 1 mm with acquisition time of 8 min, 13 s. T2-weighted images: TR = 3200 ms, TE = 402 ms, FOV = 256 mm, matrix = 258 × 256, slice thickness = 1 mm. For the twelve runs of functional images—one for each block of trials—the scans were T2*-weighted, echo-planar images with a TR = 2 s, TE = 30 ms, flip angle = 90º, 32 axial slices, voxel size = 3 × 3 × 3 mm, 4 mm slice thickness, FoV = 256 mm, with 64 × 64 grid.

#### 2.3.1. Brain regions of interest

In addition to the whole-brain cortical thickness analysis, brain regions that previously had reported associated with attentional filtering process, working memory capacity or AD-related learning/memory performance [middle frontal gyrus, basal ganglia (caudate, putamen, globus pallidus), intraparietal sulcus, medial temporal lobe (hippocampus, and entorhinal and parahippocampal cortices); Figure 2) were selected as regions-of-interest (ROIs). The ROIs were defined for each subject using Freesurfer. The segmentation quality then was confirmed visually by a reviewer blinded to group assignment. The right and left hemisphere ROI data values were averaged.

**Figure 2.**
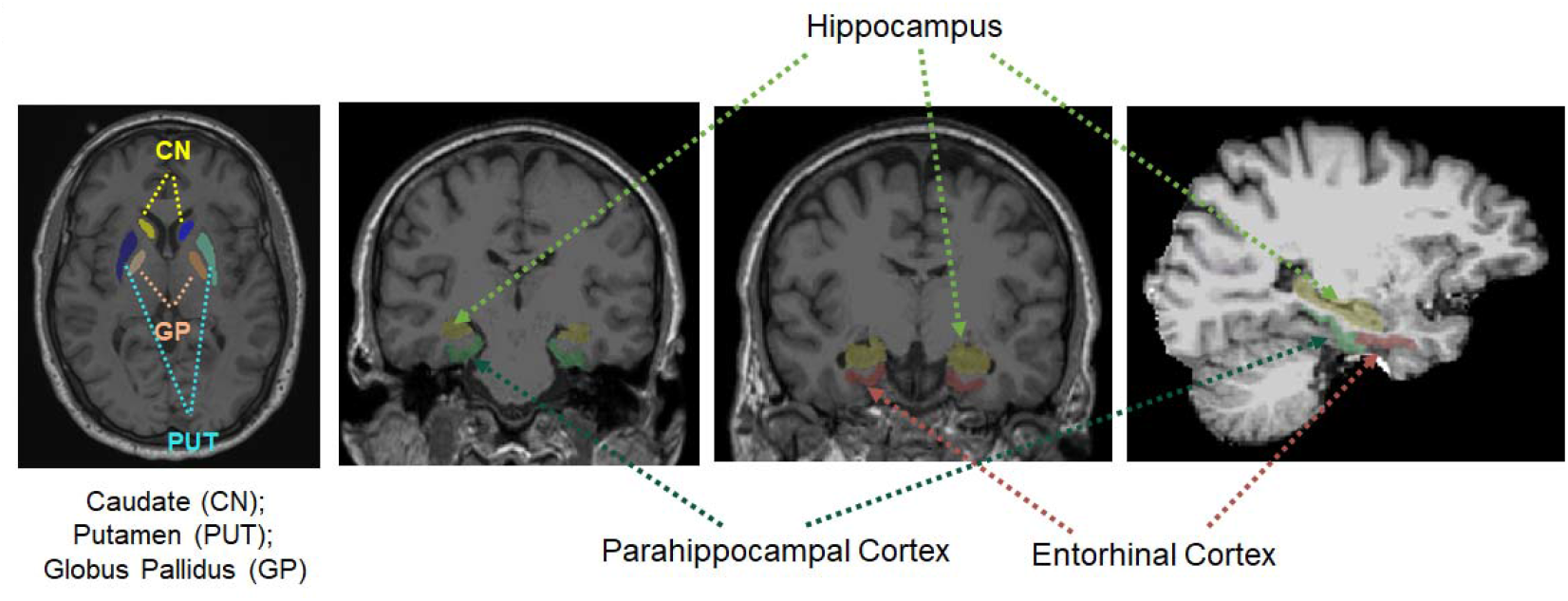
Brain regions of interests

#### 2.3.2. Volumes and thickness

Volumetric segmentation and cortical parcellation for thickness calculation were performed with the Freesurfer image analysis suite (http://surfer.nmr.mgh.harvard.edu/). The processing included motion correction, removal of non-brain tissue using a hybrid watershed/surface deformation procedure (Ségonne et al. 2004), automated Talairach transformation, and segmentation of the deep gray matter volumetric structures and parcellation of cortical gay matter structures (Fischl et al. 2002; Fischl et al. 2004).

### 2.4. Statistical Analysis

Group comparisons of demographic data were conducted using one-way analysis of variance (ANOVA) or χ^2^ test. Group comparisons of neuropsychological and MRI structural ROI metrics were conducted using multivariate analysis of covariance (MANCOVA) using age and education as covariates. When comparing the hippocampal volume, total intracranial volume (TIV) additionally was used as a covariate. The primary neuropsychological metrics were *K* scores that were derived from hit rate (proportion of correct responses when a change was present) and false alarm rate (proportion of incorrect responses on no-change trials): *K = N * (H-FA)*, where *N* is the number of relevant, to-be-stored items, *H* is the hit rate and *FA* is the false alarm rate as suggested by Cowan (2001). Association analyses of MRI structural (volume and cortical thickness) with K scores were conducted for controls and PD patients separately using Pearson partial correlation analyses with adjustment for age and education. Statistical significance was defined as α=0.05. SAS 9.4 was used for all statistical analyses.

## 3. Results

### 3.1. Demographics

There were no significant group differences in age, gender, education, MMSE, and depression scores (p’s >0.073).

**Table 1.** Demographics

|  | Age |  | Gender |  | Years of education |  | Hoehn & Yahr scale |  |
| --- | --- | --- | --- | --- | --- | --- | --- | --- |
|  | M | SD | Male | Female | M | SD | M | SD |
| Patients (N=19) | 66.16 | 8.81 | 14 | 5 | 16.63 | 3.25 | 2.03 | 0.77 |
| Controls (N=23) | 68.04 | 6.62 | 12 | 11 | 15.26 | 3.85 | - | - |

*Note.* Descriptive data for participants' demographics (age, gender, years of education, patients' Hoehn & Yahr scale, and years of disease).

### 3.2. Group comparison of memory metrics

In Experiment 1, there were significant group differences in overall memory conditions [F(3,35)=3.04, p=0.042) and in each individual memory condition [F(1,37)=5.79, p=0.021, R^2^=0.152 for Low-Load; [F(1,37)=8.33, p=0.007, R^2^=0.206 for Low-Load with Distractor; F(1,37)=5.89, p=0,020, R^2^=0.164 for High-Load; Figure 3a). The K socre in the Low-Load with distractor condition was significantly lower compared to that in Low-Load condition for PD (t=-2.81, p=0.008) but not for controls (t=-1.29, p=0.206).

**Figure 3.**
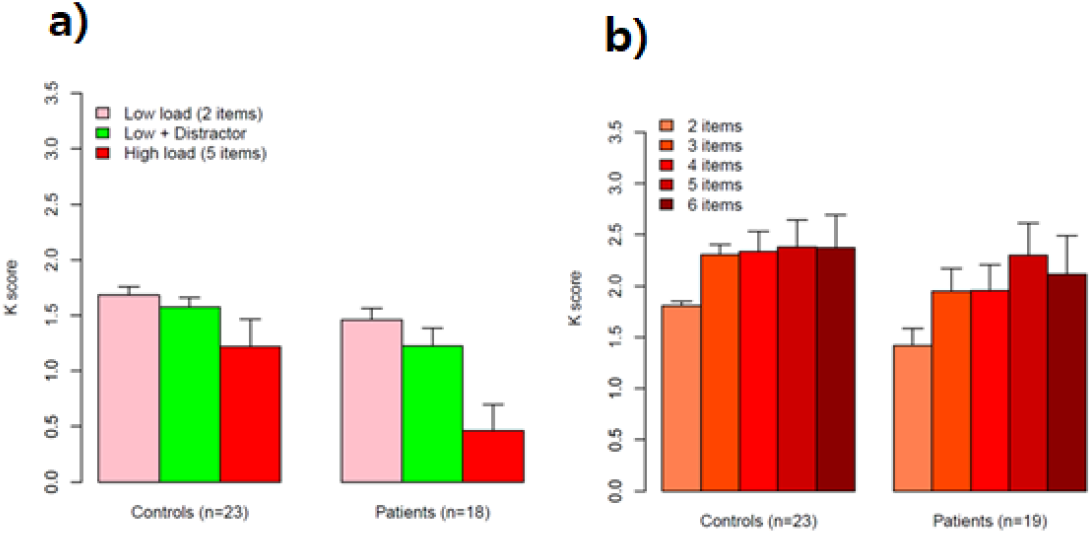
Mean K scores for controls and PD patients in 1) experiment 1 and 2) experiment 2 depending on different memory array conditions

In Experiment 2, there were no significant group differences in overall working memory condicitons (F(5,33)=0.84, p=0.534) and in each individual conditions with different memory set sizes (F’s >0.49, p’s >0.060, R^2^<0.199; Figure 3b).

### 3.3. Group comparisons in ROIs related to attentional filtering and memory storage (working memory & AD-related learning/memory storage)

There were no overall significant group differences in subcortical ROI volumes [BG and hippocampus; F(4,33)=0.69, p=0.607] or in any individual ROI volumes (F’s <1.120, p’s >=0.711). There also were no overall significant group differences in cortical ROI thicknesses [frontal gyrus, intraparietal sulci and entorhinal and parahippocampal cortices; F(3,35)=0.1.10, p=0.364] or in any individual ROI thicknesses (F’s <2.680, p’s >=0.110).

### 3.4. Associations of MRI structural metrics with memory metrics

For controls, there were no significant correlations in subcortical volumes and memory set condicitons in experiment 1 (R’s <0.380, p’s <0.099). The pallidal volume was significantly positively associated with the memory set size 2 in the experiment 2 (R=0.467, p=0.038). Volumes in other areas were not signficantly associated with any other memory set sizes in experiment 2 (R’s <0.402, p’s>0.079). There were no signficant correlations between the intraparietal cortical thickness with any memory set conditions in both Expeiment 1 and 2 (R’s <0.266, p’s >0.258)

For PD patients, among thickness metrics in individual cortical ROIs, the intraparietal cortical thickness was significantly positively associated with the memory set size 3 and 6 conditions in Experiment 2 (R=0.618, p=0.618 for set size 2 and R=0.516, p=0.034 for the set size 6) while there were no significant correlations between structural metrics and memory capacity K scores in any memory load conditions for experiment 1 (p’s >0.056, R’s <0.471). For volume metrics in individual subcortical ROIs, there were no significant correlations between any subcortical ROI volumes (BG and hippocampus) and K scores in any memory conditions in Experiment 1 and 2 (R’s <0.408, p’s >0.104).

## 4. Discussion

The present study examined nature of memory deficits and its associations with MRI structural metrics in patients with Parkinson disease (PD). Results demonstrated that, compared to controls, PD patients had lower delayed memory whereas there were no group differences in working memory scores for any memory set sizes. In addition, there were no group differences in structural metrics for any ROI and no asccociations of strucrutal metrics with delayed memory scores. The present findings suggest that both lower attentional filtering and memory storage may contribute to lower delayed memory scores in PD. Memory storage capacity for visuospatial working memory in PD, however, appeared to be comparable to that of controls at least in the absence of apparent distractors. In addition, lower delayed memory in PD may partly be associated with early brain structural or functional changes that may occur before morphological changes.

### 4.1. Lower dealyed memory due to reduced memory consolidation

Present results showed that patients’ *K* scores were comparable to those of controls when their memory was tested 2 s after the memory array offset. This result would suggest that patients with Parkinson’s disease may not have reduced storage capacity. PD patients’ ability to encode and retrieve visual information may also be comparedable to those in controls. This result is inconsistent with our previous findings demonstrating that PD patients had lower *K* scores and CDA (contralateral delay activity) amplitude, EEG correlates reflecting items held in working memory in memory conditions without distractors (Lee et al. 2010). In that experiment, howeer, a bilateral display was utilized to measure CDA, which required additional filtering by the participant even in conditions with no distracters. So it is possible that patients’ lower *K* scores and CDA amplitudes in that study could be due to impaired filtering rather than diminished storage capacity. Our current result supports this interpretation.

Instead, PD patients may have difficulty to continue holding information in memory over a prolonged period of time, 7 s, as shown in experiment 1, a period where memory consolidation process from working memory to a longer-term delayed memory may increasingly gain importance. Lower delayed memory performance in PD without working memory capacity problems may indicate reduced memory consolidation function. Altough there may be no known capacity limits for long-term memory unlike working memory that is characterized by restricted capacity limit, reduced memory consolidation ability may eventually lead to fewer items in delayed memory. It s worth mentioning that there was a paradoxical reversal from Low-Load to High-Load trials as if they were simply overwhelmed and so stopped working. This result is congruent with previous findings in which memory-load sensitive brain activation was markedly reduced when neurologically normal subjects were presented with memory loads beyond their capacity (Linden et al., 2003; Vogel & Machizawa, 2004).

### 4.2. Lower delayed memory due to impaired attentional filtering

Both controls and patients had some difficulty ignoring distracters. They demonstrated lower *K* scores compared to Low-Load condition although the number of to-be-remembered items was same for both conditions with and without distractors. Interestingly, *K* score difference between these two memory conditions was significant only for PD patients, suggesting attentional filtering difficulty in PD. This finding is consistent with previous studies reporting a critical role of basal ganlgia, especially the globus pallidus in controlling the access of incoming information into memory system (McNab and Klingberg, 2008; Cohen and Frank, 2009). Therefore, patients with basal ganglia disease might be especially vulnerable to filtering deficits. Indeed, previous studies demonstrated that even newly diagnosed, drug-naïve Parkinson’s patients showed under-activation in basal ganglia including caudate nuclei, putamen and globus pallidus when updating new information into memory system (Marklund et al., 2009). Our current finding is in line with this previous findings. The loss of dopaminergic input to the basal ganglia in Parkinson’s disease may lead to a diminished ability of the GP and reduced ability to filter out distracters so that they do not unnecessarily use up space in memory.

### 4.3. Neural correlates of lower delayed memory in PD

In the present study, we examined structural metrics (volumes and cortical thickness) in brain areas that were known to be related to attentional filtering, working memory storage, or memory consolidation processes. Consistent with comparable working memory capacity throught different memory load conditions, there were no significant group differences in the intraparietal sulcus, an area sensitie to information retention in working memory. The association analysis demonstrated that *K* scores in experiment 2 were positively associated with cortical thickness in the intraparietal sulcus, confirming the involement of this region in working memory capacity (Postle et al., 2006).

It is intriguing to note that there were no significant structural differences in any ROIs and associations between structural metrics and delayed memory scores although robustly lower delayed memory performance was obsered throught different memory conditions. It is possible that our sample size was too small reliably to detect structural differences. It also is possible that patients’ lower delayed memory observed in this study may partly be associated with early brain microstructural or functional changes that may occur before morphological changes. Further studies utilizing brain imaging markers that sensitiely capture early brain microstructural and functional changes should be warranted to confirm current findings and to elucidate neural correlates of delayed memory in PD.

## 5. Conclusions

Lower attentional filtering and memory consolidation problems into long-term memory storage may contribute to lower delayed memory performance in PD.

## Data Availability

All data produced in the present study are available upon reasonable request to the authors

## Supplementary Materials

No Supplementary Materials are available.

## Institutional Review Board Statement

All participants gave their informed consent according to procedures approved by the ethics board at the University of Missouri-Columbia (protocol code: 1170557 on 2010.09.01).

## Informed Consent Statement

Informed consent was obtained from all subjects involved in the study.

## Data Availability Statement

Data can be available upon request.

## Acknowledgments

This work was supported by the Dong-A University research fund.

